# Willingness to Vaccinate against COVID-19 among Healthcare Workers: An Online Survey in 10 Countries in the Eastern Mediterranean Region

**DOI:** 10.1101/2021.03.20.21253892

**Authors:** Yasir Ahmed Mohammed Elhadi, Azza Mehanna, Yusuff Adebayo Adebisi, Haider M. El Saeh, Saddam Abdulhakem Alnahari, Omar Hassan Alenezi, Diala El Chbib, Zahraa Yahya, Eiman Ahmed, Shoaib Ahmad, Saad Uakkas, Majdi Mohammed Sabahelzain, Bushra Ahmed Alyamani, Arash Nemat, Don Eliseo Lucero-Prisno, Ashraf Zaghloul

## Abstract

**Background:** Willingness of healthcare workers to be vaccinated is an important factor to be consider for successful COVID-19 vaccination programme. Our study aimed to understand the willingness of health workers to receive COVID-19 vaccine and associated concerns across 10 countries in the Eastern Mediterranean Region (EMRO).

**Method:** A cross-sectional study was conducted in January 2021 among healthcare workers using an online survey. A total of 2806 health workers (Physicians, Nurses and Pharmacists) completed and returned the informed consent along with the questionnaire electronically. Data were analyzed using IBM SPSS software package version 20.0. S

**Results:** More than half of the respondents (58.0%) were willing to receive COVID-19 vaccine, even if the vaccination is not mandatory for them. On the other hand, 25.7% of respondents were not willing to undertake COVID-19 vaccination while 16.3 % answered undecided. The top three reasons for not intending to be vaccinated were unreliability of COVID-19 vaccine clinical trials (62.0%), fear of the side effects of the vaccine (45.3%), and that COVID-19 vaccine will not give immunity for a long period of time (23.1%).

**Conclusion:** Overall, our study revealed suboptimal acceptance of COVID-19 vaccine among our respondents in the EMRO region. Significant refusal of COVID-19 vaccine among healthcare professionals can reverse hard-won progress in building public trust in COVID-19 vaccination program. Our findings suggest the need to develop tailored strategies to address concerns identified in the study in order to ensure optimal vaccine acceptance among healthcare workers in the EMRO.

## 1. Introduction

Coronavirus disease (COVID-19) is an infectious disease caused by the novel coronavirus that was first discovered in Wuhan, China. In 2020, the World Health Organization (WHO) declared the outbreak a global health emergency and later a pandemic after it has spread to many countries of the world (Cucinotta & Vanelli, 2020). COVID-19 pandemic has raised immense global concerns for humanity and has posed unprecedented challenges to healthcare systems worldwide. As of 1 February 2021, globally, there have been 102,399,513 confirmed cases of COVID-19 reported to the World Health Organization (WHO), including 2,217,005 deaths “WHO Coronavirus Disease”. Unsurprisingly, healthcare workers account for a number of the reported cases (Gagneux-Brunon et al., 2021).

The healthcare workers are on the first line of the battle against the COVID-19 (Nemat et al., 2020). Thus, protecting them should be one of the top priorities in the fight against COVID-19. Their contacts with patients can facilitate the spread of the virus (Jin et al., 2020). More so, healthcare workers are at highest risk of COVID-19 exposure and mortality due to work environment conditions, including personal protective equipment (PPE) shortages, insufficient staffing, and inadequate safety training and preparation amid the COVID-19 pandemic (Nemat et al., 2020). As of July 2020, the United Nations announced that over 1.4 million infections of COVID-19 are accounted for in healthcare workers, at least 10% of all cases “Coronavirus latest”. Comparisons of healthcare workers with and without COVID-19 infection showed an increased relative risk related to personal protective equipment, workplace setting, profession, exposure, contacts, and testing (Gholami et al., 2021).

In low-income countries, healthcare workers are more vulnerable to the devastating impacts of COVID-19. Being away from home and facing the hardship of fighting COVID-19 put healthcare workers in dire situation (Nemat et al., 2020). Moreover, the economic hardship is another major problem healthcare workers facing amid the pandemic (Kelley et al., 2020). The COVID-19 pandemic continues to pose multiple health challenges across the world. In the Eastern Mediterranean Region where morbidity and mortality from the disease remain a serious cause for concern (Al-Mandhari, 2020). Concern like this can pose serious psychological health impacts on healthcare workers (Lai et al., 2020), hence causing serious health implications for them.

Since the emergence of the pandemic, the world is desperately waiting for a safe and effective vaccine. Efforts such as preventative measures have been put into action to curb the spread of the virus (Koirala et al., 2020). However, implementing a global vaccination program with broad range of clinical and socioeconomic benefits is the most effective mean to end the pandemic (Adebisi, Alaran, et al., 2020). As of December 2020, several vaccines against COVID-19 have been authorized (Schaffer Deroo et al., 2020). However, with the growing vaccination coverage, worldwide, under-vaccinated or non-vaccinated communities are still a concern for vaccination programs (Pugliese-Garcia et al., 2018).

The complex phenomenon of vaccine hesitancy refers to “delay in acceptance or refusal of vaccines despite availability of vaccine services” (Dubé et al., 2014), it is one of the top public health issues listed by WHO (Graham, 2019). Despite of proven safety, efficacy and effectiveness of vaccines, an increasing number of individuals perceive vaccines as unsafe and unnecessary (Dubé et al., 2013). Addressing vaccine hesitancy among healthcare workers is crucial. Healthcare workers are considered one of the most important strata of society and a priority target group for COVID-19; vaccinating them is an utmost task for the world. However, intention to take COVID-19 vaccine depends on the confidence and safety of the vaccine. Amid the pandemic, healthcare workers have also shown skepticism towards vaccine even in developed countries (Dror et al., 2020; Karafillakis et al., 2016). This is a point of great concern for the world since healthcare workers are the most credible and trusted sources of the information and their doubt on vaccine will subjugate other people to follow the same pattern of believe.

To find out the notion of healthcare workers towards vaccination in a bigger context, a study is required. This research is aimed at understanding the willingness of healthcare workers (Physicians, Nurses, and Pharmacists) to receive COVID-19 vaccination from 10 countries in Eastern Mediterranean Region.

### 2. Method

### 2.1 Study Design and Sampling Technique

This is a cross-sectional survey among health workers (physicians, nurses, and pharmacists) in 10 countries in Eastern Mediterranean region (Afghanistan, Egypt, Iraq, Kuwait, Lebanon, Libya, Morocco, Pakistan, Sudan, and Yemen). Non-probability convenient sampling technique was used to recruit the respondents. The inclusion criteria were being a physician, nurse or pharmacist working in one of targeted countries at the time of data collection and having access to an internet connection to fill out the online questionnaire. Individuals who do not consent to participate in the study were excluded.

### 2.2 Study Instrument and Administration

A short online questionnaire was developed after a review of similar study (Adebisi, Alaran, et al., 2020) and it comprises sections on the demographic characteristics of the respondents including; nationality, age, sex, marital status, profession and years of working experience (independent variables).

Outcome variables include the respondents’ willingness to vaccinate against COVID-19 and the reasons not willing to undertake COVID-19 vaccination among non-intenders. Question 1: if you are given the option to choose to take COVID-19 vaccine, will you take the vaccine? (With a Yes, No or Undecided options). Participants not willing or undecided yet were further asked question 2: what is your reason(s) for not accepting COVID19 vaccine uptake?

The questions were entered into an online survey system and a link to the electronic questionnaire was generated. The final questionnaire was distributed by the research team members in each country to respondents across social media platforms, specifically groups of health workers as used in previous studies (Awucha et al., 2020). The data collected took place in January 2021.

### 2.3 Data Analysis

Data were fed to the computer and analyzed using IBM SPSS software package version 20.0 (Armonk, NY: IBM Corp). The Kolmogorov-Smirnov was used to verify the normality of distribution of variables and comparisons between groups for categorical variables were assessed using Chi-square test. Binary logistic regression analysis was carried out to identify parameters more strongly associated with respondents’ willingness to vaccinate against COVID-19. Significance of the obtained results was judged at the 5% level.

### 2.4 Ethical Considerations

This study has been approved by Ethics and Technical Committee of High Institute of Public Health Alexandria University. Confidentiality and anonymity were also ensured by not putting names or attaching any identifiable codes to the online questionnaires, and the rights of the participants to withdraw anytime from the study were also clearly stated in the online survey.

## 3. Results

Distribution and sociodemographic characteristics of respondents: A total of 2806 health workers representing females (50.4%) and males (49.6%) from 10 countries in the EMRO completed the online survey. The mean age of participants was 31.3 (± 9.1 years). Most respondents were physicians (58.0%). Table 1 shows the distribution and sociodemographic characteristics of studied samples.

**Table 1.**
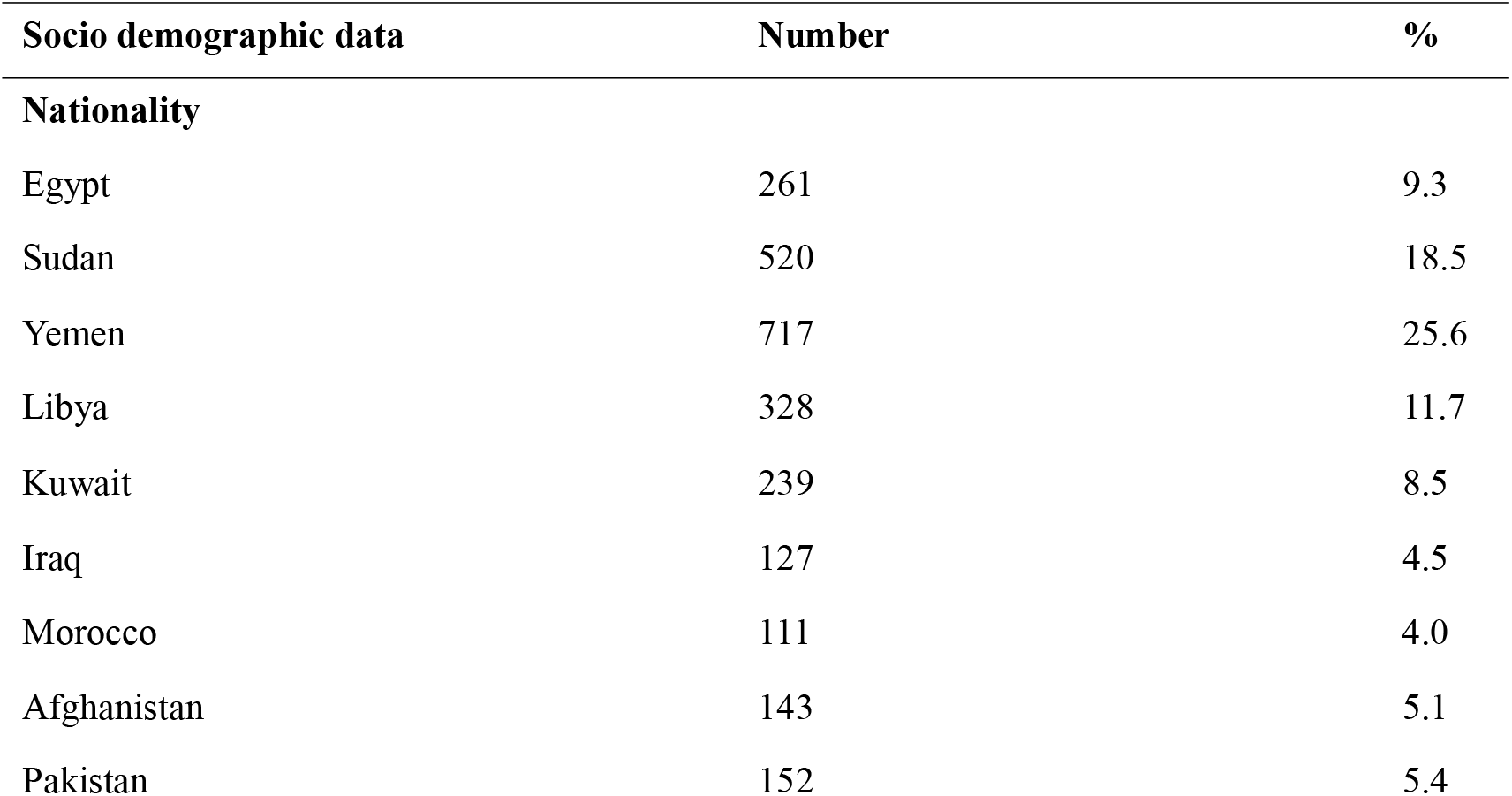
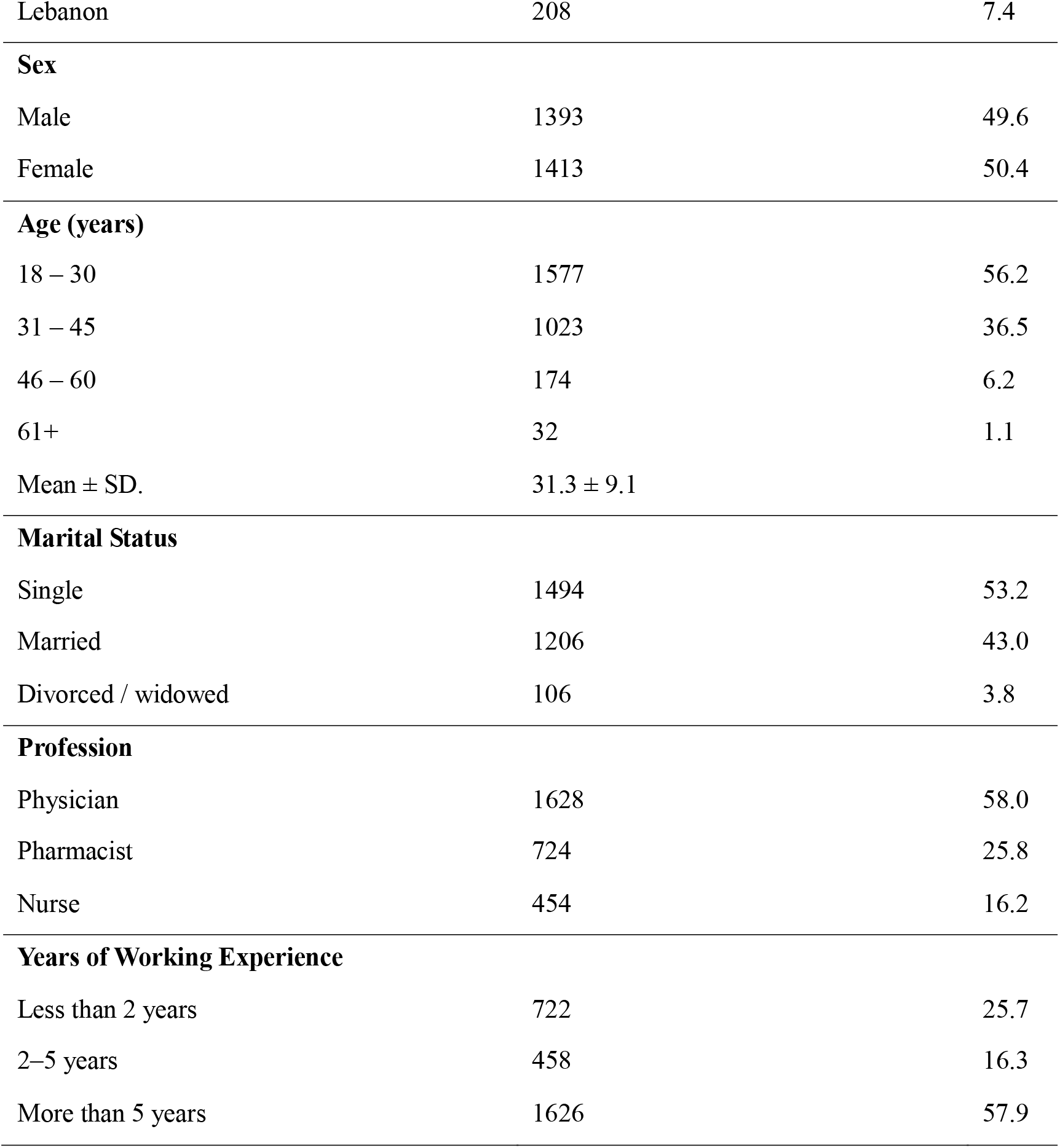
Distribution and Sociodemographic Characteristics of Respondents.

|  |  |  |
| --- | --- | --- |
| Socio demographic data | Number | % |
| **Nationality** |  |  |
| Egypt | 261 | 9.3 |
| Sudan | 520 | 18.5 |
| Yemen | 717 | 25.6 |
| Libya | 328 | 11.7 |
| Kuwait | 239 | 8.5 |
| Iraq | 127 | 4.5 |
| Morocco | 111 | 4.0 |
| Afghanistan | 143 | 5.1 |
| Pakistan | 152 | 5.4 |
| Lebanon | 208 | 7.4 |
| **Sex** |  |  |
| Male | 1393 | 49.6 |
| Female | 1413 | 50.4 |
| **Age (years)** |  |  |
| 18 – 30 | 1577 | 56.2 |
| 31 – 45 | 1023 | 36.5 |
| 46 – 60 | 174 | 6.2 |
| 61+ | 32 | 1.1 |
| Mean ± SD. | 31.3 ± 9.1 |  |
| **Marital Status** |  |  |
| Single | 1494 | 53.2 |
| Married | 1206 | 43.0 |
| Divorced / widowed | 106 | 3.8 |
| **Profession** |  |  |
| Physician | 1628 | 58.0 |
| Pharmacist | 724 | 25.8 |
| Nurse | 454 | 16.2 |
| **Years of Working Experience** |  |  |
| Less than 2 years | 722 | 25.7 |
| 2–5 years | 458 | 16.3 |
| More than 5 years | 1626 | 57.9 |

### 3.1 Health workers’ willingness to vaccinate against COVID-19

More than half of the respondents (58.0%) were willing to receive COVID-19 vaccine, even if the vaccination is not mandatory for them. On the other hand, 25.7% of respondents were not willing to take COVID-19 vaccine while 16.3% answered undecided. Figure 1 shows willingness to vaccinate against COVID-19 among health professional in studied countries.

**Figure 1:**
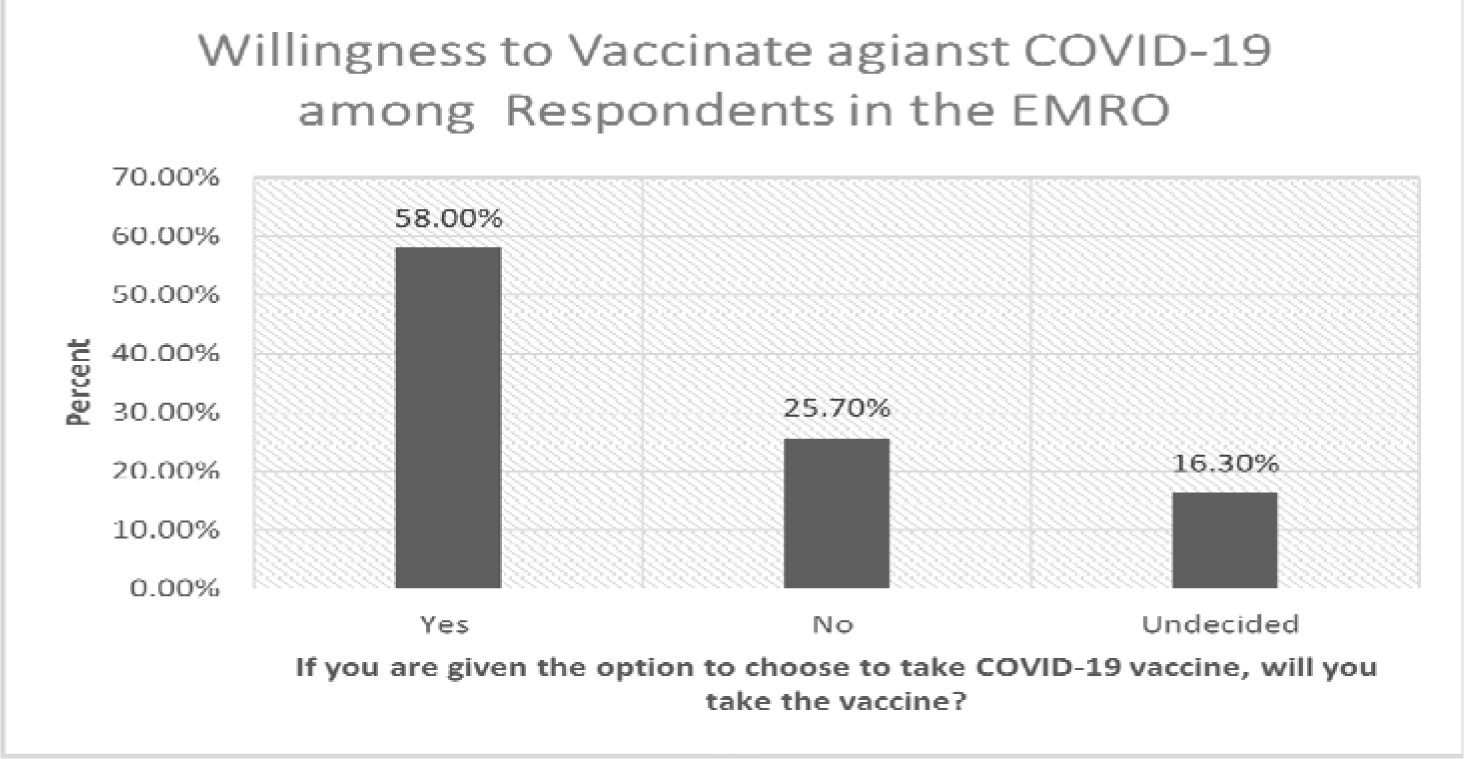
Willingness to vaccinate against COVID-19 among health professional in studied countries.

### 3.2 Variables Associated with Respondents’ Intention to Receive COVID-19 Vaccination

As shown in Table 2 results indicated respondent’s nationality (P value <0.001), sex (P value <0.001), marital Status (P value <0.022), profession (P value <0.001) and years of working experience were significantly associated with willingness to receive COVID-19 vaccination. However, there was no significant association between age group and willingness to receive COVID-19 vaccine. Additionally, Figure 2 below shows high vaccine acceptance among healthcare professionals working in Afghanistan (72.0%) and Libya (70.0%).

**Table 2.** Association between sociodemographic variables and respondents’ willingness to receive COVID-19 Vaccine.

| Sociodemographic variables | If you are given the option to choose to take COVID-19 vaccine, will you take the vaccine? |  | Test of Significance |  |
| --- | --- | --- | --- | --- |
| | Yes (n=1626) | No/Undecided (n=1180) | $\chi^2$ | p |
| Nationality |  |  |  |  |
| Egypt | 119 (7.3%) | 142 (12.0%) | 122.43* | <0.001* |
| Sudan | 292 (18.0%) | 228 (19.3%) |  |  |
| Yemen | 464 (28.5%) | 253 (21.4%) |  |  |
| Libya | 230 (14.1%) | 98 (8.3%) |  |  |
| Kuwait | 103 (6.3%) | 136 (11.5%) |  |  |
| Iraq | 58 (3.6%) | 69 (5.8%) |  |  |
| Morocco | 64 (3.9%) | 47 (4.0%) |  |  |
| Afghanistan | 103 (6.3%) | 40 (3.4%) |  |  |
| Pakistan | 106 (6.5%) | 46 (3.9%) |  |  |
| Lebanon | 87 (5.4%) | 121 (10.3%) |  |  |
| Sex |  |  |  |  |
| Male | 926 (56.9%) | 467 (39.6%) | 82.55* | <0.001* |
| Female | 700 (43.1%) | 713 (60.4%) |  |  |
| **Age (years)** |  |  |  |  |
| 18 – 30 | 904 (55.6%) | 673 (57%) | 1.766 | 0.622 |
| 31 – 45 | 608 (37.4%) | 415 (35.2%) |  |  |
| 46 – 60 | 96 (5.9%) | 78 (6.6%) |  |  |
| 61+ | 18 (1.1%) | 14 (1.2%) |  |  |
| **Marital Status** |  |  |  |  |
| Single | 865 (53.2%) | 629 (53.3%) | 7.66* | 0.022* |
| Married | 713 (43.8%) | 493 (41.8%) |  |  |
| Divorced / widowed | 48 (3%) | 58 (4.9%) |  |  |
| **Profession** |  |  |  |  |
| Physicians | 993 (61.1%) | 635 (53.8%) | 20.69* | <0.001* |
| Pharmacists | 369 (22.7%) | 355 (30.1%) |  |  |
| Nurses | 264 (16.2%) | 190 (16.1%) |  |  |
| **Experience (Years)** |  |  |  |  |
| Less than 2 years | 0 (0%) | 722 (61.2%) | 2806.0* | <0.001* |
| 2–5 years | 0 (0%) | 458 (38.8%) |  |  |
| More than 5 years | 1626 (100%) | 0 (0%) |  |  |

χ2: Chi square test
\*: Statistically significant at $p \leq 0.05$

**Figure (2):**
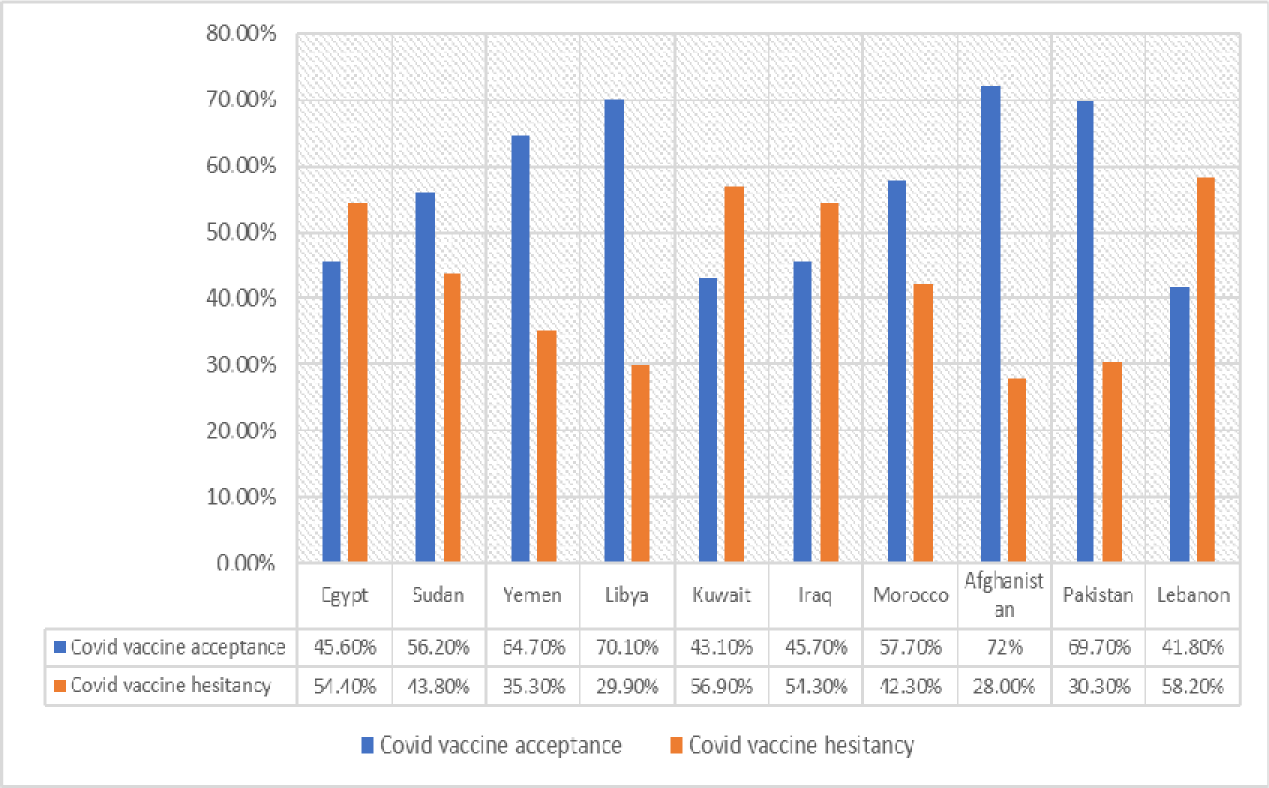
COVID-19 vaccine acceptance and hesitancy among health workers by countries.

### 3.3 Parameters Affecting Health Workers’ Willingness to Receive COVID-19 vaccination

As shown in Table 3. Health professionals working in; Sudan (P <0.001); Yemen (P <0.001); Libya (P <0.001); Afghanistan (P <0.001); Pakistan (P <0.001) or Morocco (P <0.001), were more likely to be willing to receive COVID-19 vaccine than health professionals working in the other countries. However, females (P <0.001), pharmacists (P <0.001) and older ages (P <0.001) were more significantly associated with having reservations towards vaccination (not willing or didn’t decide to receive COVID-19 vaccine).

**Table 3:** Multivariate binary logistic regression analysis for the parameters affecting health workers’ willingness to receive COVID-19 vaccination.

| Variables | p | OR | 95% C.I |  |
| --- | --- | --- | --- | --- |
|  |  |  | UL | LL |
| Nationality |  |  |  |  |
| Egypt | 0.308 | 0.820 | 0.560 | 1.201 |
| Sudan | <0.001* | 0.541 | 0.388 | 0.754 |
| Yemen | <0.001* | 0.408 | 0.293 | 0.568 |
| Libya | <0.001* | 0.305 | 0.208 | 0.449 |
| Kuwait | 0.965 | 0.991 | 0.666 | 1.474 |
| Iraq | 0.967 | 0.991 | 0.628 | 1.563 |
| Morocco | 0.019* | 0.566 | 0.352 | 0.912 |
| Afghanistan | <0.001* | 0.364 | 0.227 | 0.584 |
| Pakistan | <0.001* | 0.397 | 0.251 | 0.628 |
| Lebanon | 0.308 | 0.820 | 0.560 | 1.201 |
| **Age (Years)** | 0.031* | 1.013 | 1.001 | 1.024 |
| **Sex (Female)** | <0.001* | 1.898 | 1.610 | 2.238 |
| **Marital Status** |  |  |  |  |
| Single® |  |  |  |  |
| Married | 1.039 | 0.843 | 1.281 | 1.039 |
| Divorced / widowed | 1.473 | 0.950 | 2.283 | 1.473 |
| **Profession** |  |  |  |  |
| Physician® |  |  |  |  |
| Pharmacist | <0.001* | 1.422 | 1.171 | 1.725 |
| Nurse | 0.647 | 0.947 | 0.751 | 1.195 |

OR: Odds ratio, CI: Confidence interval , LL: Lower limit, UL: Upper Limit, ®: reference group, \*: Statistically significant at p ≤ 0.05

### 3.4 Vaccination Hesitancy among Health Workers in the Studied EMRO

Participants not willing or did not decide to take COVID-19 vaccine, despite of vaccine availability, confirmed safety and efficacy were regarded as “being hesitant to receive COVID-19 vaccine”. Figure 2 below shows the COVID-19 vaccine acceptance, and hesitancy level among health workers by countries, with the highest vaccine hesitancy observed in Lebanon (58.2%), Kuwait (56.9%) and Egypt (54.4%).

### 3.5 Reasons Associated with COVID-19 Vaccination Hesitancy

Table 4 shows different reasons associated with COVID-19 vaccine hesitancy among our respondents. The unreliability of COVID-19 vaccine clinical trials and fear of the vaccine’s side effects were reported as reasons behind COVID-19 vaccine hesitancy by 62.0% and 45.0% of the study participants, respectively. Approximately 23.0% of the healthcare professionals thought that the vaccine would not give immunity for an extended period.

**Table 4.**
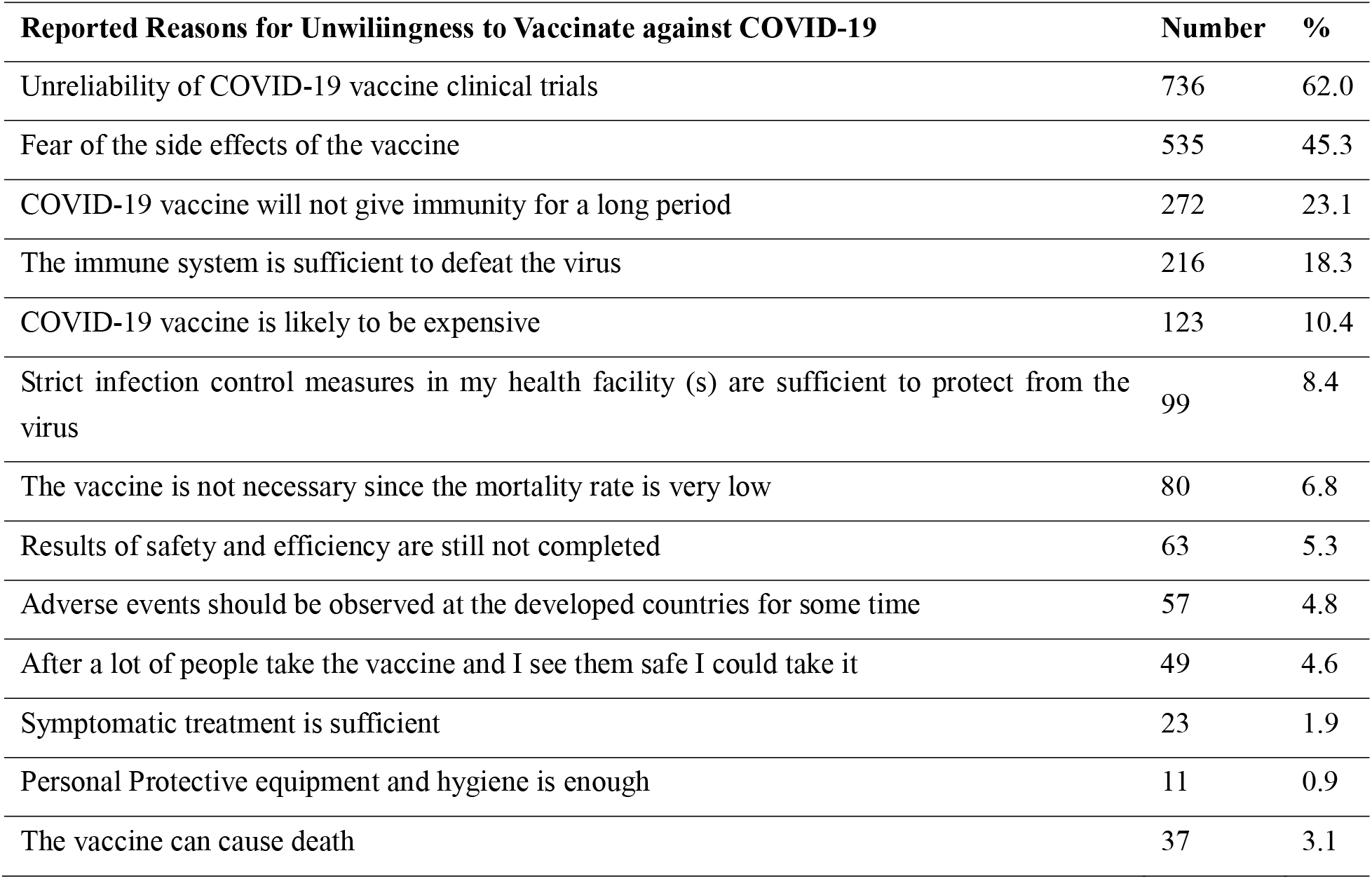
Respondents Concerns Towards COVID-19 vaccination.

| Reported Reasons for Unwillingness to Vaccinate against COVID-19 | Number | % |
| --- | --- | --- |
| Unreliability of COVID-19 vaccine clinical trials | 736 | 62.0 |
| Fear of the side effects of the vaccine | 535 | 45.3 |
| COVID-19 vaccine will not give immunity for a long period | 272 | 23.1 |
| The immune system is sufficient to defeat the virus | 216 | 18.3 |
| COVID-19 vaccine is likely to be expensive | 123 | 10.4 |
| Strict infection control measures in my health facility (s) are sufficient to protect from the virus | 99 | 8.4 |
| The vaccine is not necessary since the mortality rate is very low | 80 | 6.8 |
| Results of safety and efficiency are still not completed | 63 | 5.3 |
| Adverse events should be observed at the developed countries for some time | 57 | 4.8 |
| After a lot of people take the vaccine and I see them safe I could take it | 49 | 4.6 |
| Symptomatic treatment is sufficient | 23 | 1.9 |
| Personal Protective equipment and hygiene is enough | 11 | 0.9 |
| The vaccine can cause death | 37 | 3.1 |

## 4. Discussion

Vaccination is an important public health tool and one of the most important advances in healthcare in the fight against infectious diseases (Alaran et al., 2021). It is responsible for the eradication of rinderpest and smallpox and the control of infectious diseases such as polio in many parts of the world (Adebisi, Eliseo-Lucero Prisno, et al., 2020). It is therefore clear that a safe, highly effective, and globally acceptable and equitable vaccination program, together with pre-existing precautionary measures, is essential to effectively contain the outbreak (Lucero-Prisno et al., 2021).

It is often mistakenly believed that healthcare professionals attitudes must be positive towards vaccines because they have scientific and medical training. However, this is not always true because healthcare workers are not a homogenous group, and most are not experts in the field of vaccination. Our study present insights regarding the willingness of healthcare professional in the EMRO region. Healthcare workers are among the priority group to receive vaccination, so it is important to understand their willingness to take COVID-19 vaccine or not (Shekhar et al., 2021). This will better provide insights to address barriers to widespread COVID-19 vaccination acceptance.

Interestingly, we found that overall, more than half (58.0%) of our respondents are willing to take the COVID-19 vaccine. Our study also revealed that less than half of our respondents in Egypt, Kuwait, Iraq, and Lebanon are willing to take the COVID-19 vaccine. Similarly, a study also revealed that 28% of healthcare workers in Democratic Republic of Congo are willing to take the COVID-19 vaccine (Ditekemena et al., 2021). A study among healthcare workers in the United States revealed that only one in three said they would take the vaccine immediately it becomes available (Shekhar et al., 2021). More so, in Malta, only half of the participants (healthcare workers) stated that they intend to take the COVID-19 vaccine (Grech et al., 2020). Additionally, less than half of the healthcare workers in a survey conducted in France and French-speaking part of Belgium and Canada showed high acceptance of COVID-19 vaccine (Verger et al., 2021). This is worrisome in that healthcare professionals are expected to have considerate believe in the safety and efficacy as well as the enormous benefits COVID-19 vaccine can reap for the pandemic response. In addition, healthcare workers, as models, are typically entrusted with the task of providing reliable information regarding health issues to the public and this is associated with greater compliance with health interventions. Furthermore, our study revealed that high vaccine acceptance among healthcare professionals working in Afghanistan (72.0%) and Libya (70.0%). This can be compared to a study in Turkey, where 68.8% of the healthcare worker are willing to take the COVID-19 vaccine (Kose et al., 2021).

Findings from our study also revealed that the top three reasons for not willing to be vaccinated are “unreliability of COVID-19 vaccine clinical trials”, “fear of the side effects of the vaccine”, and “COVID-19 vaccine will not give immunity for a long period.” This is worrisome in that healthcare professional’s recommendations play a key role in their patients’ vaccination behavior. They serve as an important source of information for the general public and their consultation can also be a key factor in patients’ decision to be vaccinated or not. There is a significant need to address concerns and increase awareness to improve chances for higher acceptance of a COVID-19 vaccine. Otherwise, this poses additional possibility of mass rejection of COVID-19 vaccine in the general population.

A major strength of our study is the large sample size. In addition, our survey population is also diverse with representation from different genders, age groups, ethnic and roles in healthcare. However, our study is not without limitations. There is a risk of selection bias, and this would limit the generalizability of our findings to all the healthcare professionals in the EMRO. Additionally, our study only included nurses, physicians, and pharmacists. Despite these limitations, these findings are not inconsistent with the findings from previous studies about willingness to be vaccinated or vaccine hesitancy among healthcare workers. The survey questionnaire was available in English and distributed in an online format, which can further introduce selection bias favoring English-literate HCWs and those with access to the Internet. Social desirability bias may also affect the interpretation of our study results. Most importantly, the survey was conducted in January 2021 when information regarding COVID-19 vaccines may have not circulated widely. Therefore, it possible that intention to be vaccinated would have changed.

## 5. Conclusion

Overall, our study revealed suboptimal acceptance of COVID-19 vaccine among our respondents in the EMRO region. Significant refusal of COVID-19 vaccine among healthcare professionals can reverse hard-won progress in building public trust in COVID-19 vaccination program. Our findings suggest the need to develop tailored strategies to address concerns identified in the study in order to ensure optimal vaccine acceptance among healthcare workers in the EMRO.

## Data Availability

The data presented in this study are available on request from the corresponding author.

## Acknowledgments

We would like to thank Mohammed Yasir Essar for his assistance in conducting the study.

## Informed Consent Statement

Informed consent was obtained from all subjects involved in the study.

## Data Availability Statement

The data presented in this study are available on request from the corresponding author.

